# The effects of COVID-19 on European healthcare provision for people with major depressive disorder: a scoping review protocol

**DOI:** 10.1101/2022.02.17.22269638

**Authors:** Dilveer Sually, Win Lee Edwin Wong, Diego Hidalgo-Mazzei, Vinciane Quoidbach, Judit Simon, Patrice Boyer, Rebecca Strawbridge, Allan H. Young

## Abstract

Even before the pandemic, the treatment gaps in depression care were substantial, with issues ranging from rates of depression detection and intervention to a lack of follow-up after treatment initiation and access to secondary care services. The COVID-19 pandemic, which has had major effects on global healthcare systems, is almost certain to have impacted the MDD care pathway, though it is unclear what changes have manifested and what opportunities have arisen in response to COVID-19. The extent to which patients receive best-practice care is likely closely linked to clinical outcomes (and therefore disability burden) and as such, it is important to examine treatment gaps on the MDD care pathway during the pandemic. Here, we outline a protocol for a scoping review that investigates this broad topic, focusing on continuity of care and novel methods (e.g. digital approaches) used to mitigate care disruption. This scoping review protocol was designed according to the Preferred Reporting Items for Systematic Reviews and Meta-Analyses extension for scoping reviews (PRISMA-ScR) standards and will culminate in a narrative synthesis of evidence.

## Introduction

The burden of mental health problems in Europe has long been substantial, with data from 2018 indicating that over one in six people across the EU population is affected by a mental health disorder in a year [1] translating to substantial costs in terms of suffering and economics. We previously synthesized data from across Europe on the extent of ‘treatment gaps’ in the care of people with major depressive disorder (MDD), finding substantial discrepancies between best practice and current practice spanning entire care pathways for MDD [2, 3].

The COVID-19 pandemic has impacted the healthcare sector to an unprecedented extent, with more than 300 million confirmed cases and 5.5 million deaths [4]. Although COVID-19 has clearly impacted mental health care pathways [5, 6, 7], the precise nature of these effects is unknown, and this is a nuanced and multifaceted topic. Continuity of care in mental health is itself a complex concept, with some ambiguity on the key elements of continuity, though much work has been done to clarify this concept [8, 9]. Clearly the pandemic has exacerbated healthcare inequalities [10] and increased the prevalence of adverse psychiatric symptoms [11], yet it has also been suggested that COVID-19 presents an opportunity to reset fragmented health and care systems [12, 13], a chance to redesign health systems so that they are integrated, driven by people and communities, and resilient in the face of future systemic shocks. It is therefore crucial to assess how COVID-19 has changed mental health care.

Here we outline the protocol for a scoping review that builds on our previous work to indicate the effects of the pandemic on service provision for MDD in Europe. We focus on pre-specified ‘gaps’ between current and best practice spanning a stepped-care pathway as recommended by treatment guidelines, from initial detection/diagnosis through to continuity of care, specialist treatment and follow-up where required. Furthermore, this scoping review aims to synthesise data pertaining to strategies that have been implemented for MDD care during this period, as well as the impacts of these strategies. It is expected that these strategies will consist predominantly of digital or electronic health tools, which even prior to the pandemic were being introduced in attempts to enhance care across the health spectrum.

## Methods

### Design

This scoping review will be conducted in accordance with the Preferred Reporting Items for Systematic Reviews and Meta-Analyses extension for scoping reviews (PRISMA-ScR) statement [14]. The PRISMA-ScR statement was developed in an effort to standardise the greatly heterogenous nature of current scoping review methodology, and comprises a checklist of 22 reporting items, which was developed by an expert panel following recommendations from the Enhancing the QUAlity and Transparency Of health Research (EQUATOR) Network.

The stages of this review are as follows: (1) defining the key review questions and scope of review, (2) searching and screening studies that meet eligibility criteria based on title and abstracts, (3) identifying relevant studies by screening full texts, (4) extracting and evaluating data, (5) constructing a narrative synthesis to summarise results.

This review aims to build on our previous work on treatment gaps in MDD and investigate broadly what impacts the COVID-19 pandemic has had on MDD care in Europe (defined inclusively), culminating in the following specific review questions:

1. How has the pandemic affected the initiation and continuity of care (defined broadly and according to pre-specified treatment gap outcomes from our previous report) specifically for MDD?
2. What measures (particularly digital and electronic health tools) have been trialled or implemented in attempts to mitigate disruption of care for people with MDD and what impact have these had?

### Eligibility Criteria

Detailed inclusion and exclusion criteria were developed following discussion between authors. Original studies will be included in this review if they meet the following criteria:

a. Data was collected during the COVID-19 pandemic. Although timelines have varied between countries, we will take an inclusive approach, including studies from January 2020 to the date of our search.
b. Study was conducted in Europe (defined inclusively)
c. Study adopted either an interventional or observational design, and we also chose to include relevant systematic reviews (with or without a meta-analysis). We will exclude non-systematic reviews, editorial or opinion articles, and preclinical studies.
d. Study examined people with MDD (full or sub-sample), diagnosed either at baseline or during the study assessment period. A valid diagnosis of MDD should be established according to internationally recognized diagnostic criteria (e.g., ICD-10, DSM-5), which may have been recorded by a healthcare professional and/or rated using a structured or semi-structured diagnostic interview according to these diagnostic criteria.
e. Study examined working age adults (due to variations in services for young and elderly people), excluding studies confined to young people (under 18) or older people (usually aged over 65).
f. Study can be written in any language, provided that the review team are able to obtain a translation of sufficient quality within the timescales for this project.
g. Study examined one or more of our pre-specified outcomes (relating to care pathways / continuity of care, strategies for maintaining or improving care for people with depression during the COVID-19 pandemic). These outcomes are detailed further in the below section entitled “Outcome measures”.

We will include both peer-reviewed articles that report sufficient data and meeting abstracts if we are able to obtain sufficient information required for determining eligibility for inclusion in the review and synthesise results (either from the abstract itself or by contacting authors). The reference lists of relevant articles will also also hand-searched for additional articles. Authors will be electronically contacted on at least two occasions to provide necessary data if it is not available in the abstracts or manuscripts.

### Search strategy

We will search the MEDLINE (PubMed) and APA PsycINFO (via OVID) databases from 1^st^ January 2020 until the search date, inclusive, using search strings that are relevant to the COVID-19 pandemic and MDD and searching within record titles and abstracts. The following search strategy, which searches for the relevant strings within all fields will be employed for the PubMed search (and adapted for APA PsycInfo) [all fields]:

*((COVID-19 or COVID 2019 or severe acute respiratory syndrome coronavirus 2 or 2019 nCoV or SARS-COV2 or 2019nCoV or novel coronavirus) AND (major depression or major depressive disorder or major depressive episode or MDD or MDE))*

### Methodological quality assessment

Since this is a scoping review, we will not formally rate the methodological quality of included studies.

### Screening & data extraction

All studies will be imported into EndNote reference manager software, after which duplicates will be automatically removed using EndNote’s duplicate identification tool. One investigator (DS or WLEW) will screen each of the references for eligibility, initially at the title/abstract level and then at the full-text level. Any doubts that arise in this process will be resolved through consensus or discussion with another reviewer.

One author will develop an Excel data sheet for data extraction of eligible articles. Bibliographic information, such as authorship details and publication date will firstly be extracted. Subsequently, details of study characteristics will be extracted, including details on study location, study design, and objectives. Outcome data will also be extracted, as detailed further in the section entitled “Outcome measures”. Any doubts or uncertainties will be resolved through consensus or through discussion with a further reviewer.

### Outcome measures

Aligning with the review aims and objectives of this scoping review, the outcome measures pertain firstly to the pre-specified treatment gaps and secondly to strategies implemented to mitigate disruption to care.

#### Pre-specified treatment gaps

These ‘treatment gap’ outcomes are focused on the initiation and continuity of care [8], aligning with our previous European Brain Council (EBC) Value of Treatment in MDD care pathway analyses, which examined treatment gaps prior to the pandemic [2, 3]. This will therefore enable a preliminary, indicative comparison between pre-2020 and 2020-2022, to estimate the impacts of the pandemic. These outcomes are delineated in Table 1.

**Table 1.**
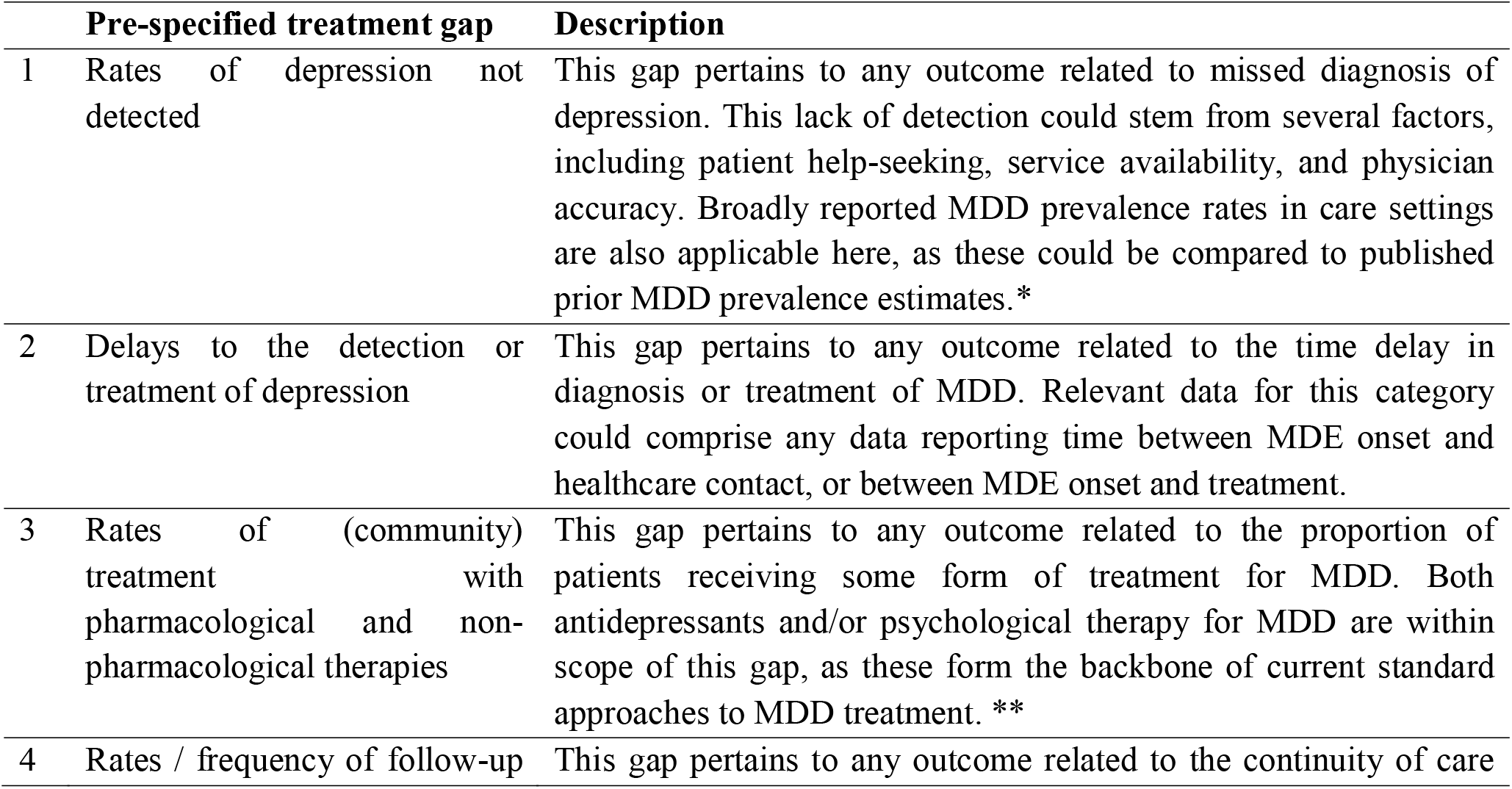
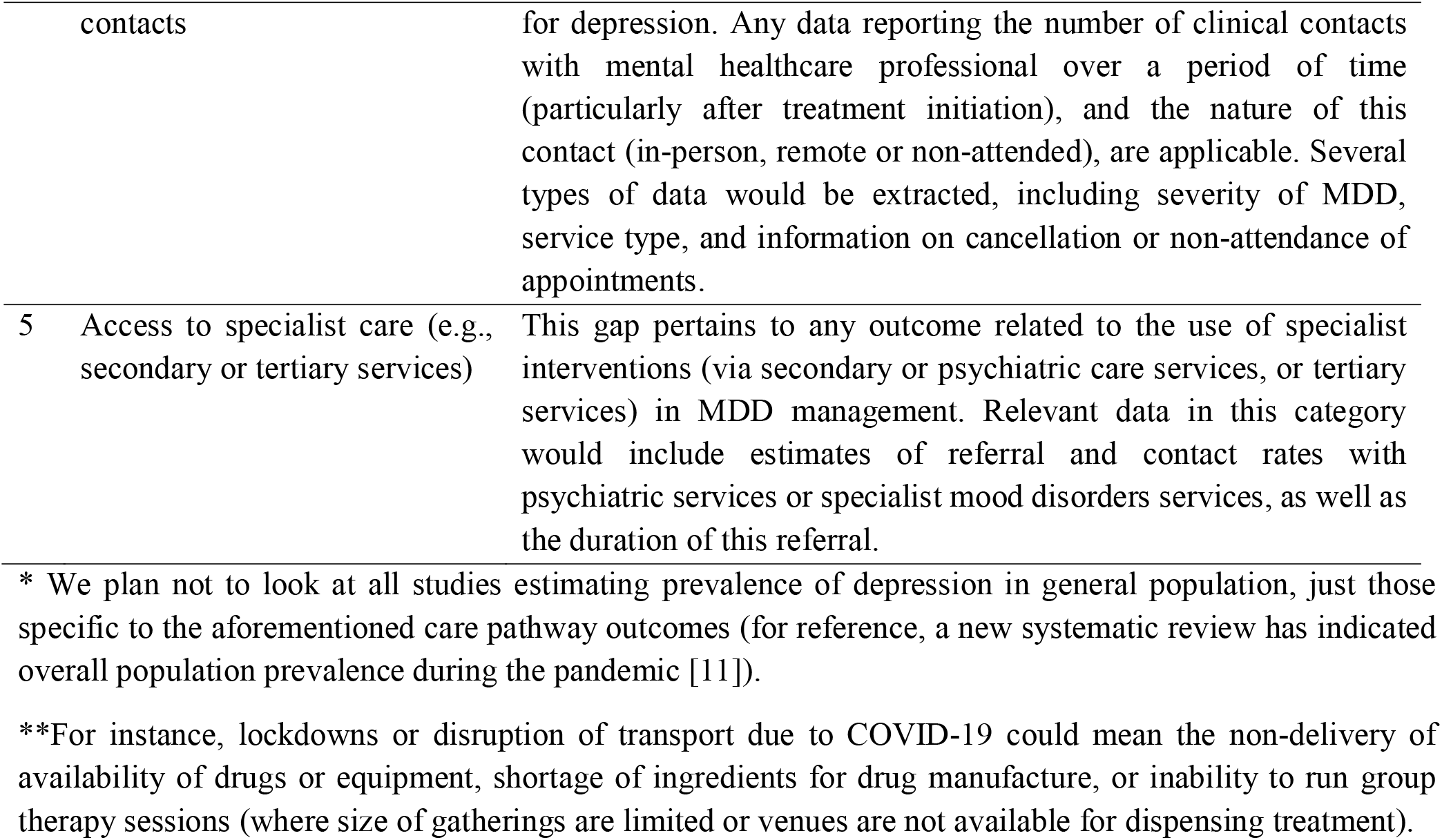

#### Mitigation strategies

These outcomes are considered more broadly; any strategies trialled/implemented in a healthcare service aiming to maintain/improve MDD care can be considered in this review.

### Strategy for data synthesis

We will summarize study characteristics and results with tables. If appropriate, figures and graphs may be constructed to illustrate linked themes that arise from the findings of this scoping review. A narrative synthesis will be constructed to explore patterns and themes that arise from the included studies; this synthesis will incorporate simple statistical process (e.g. impacts in terms of raw data and percentage metrics reported) and critical observation of trends (e.g. observing trends from graphical presentation of data as per Patel et al. [15]).

## Data Availability

All data produced in this study will be available upon reasonable request to the authors.

## Data availability

All data produced in this study will be available upon reasonable request to the authors.

## Competing interests

RS declares an honorarium from Lundbeck. AY declares the following competing interests: employed by King’s College London; Employed by King’s College London; Honorary Consultant SLaM (NHS UK). Deputy Editor, BJPsych Open. Paid lectures and advisory boards for the following companies with drugs used in affective and related disorders: Astrazenaca, Eli Lilly, Lundbeck, Sunovion, Servier, Livanova, Janssen, Allegan, Bionomics, Sumitomo Dainippon Pharma, COMPASS, Sage, Novartis. Consultant to Johnson & Johnson. Consultant to Livanova. Received honoraria for attending advisory boards and presenting talks at meetings organised by LivaNova. Principal Investigator in the Restore-Life VNS registry study funded by LivaNova. Principal Investigator on ESKETINTRD3004: “An Open-label, Long-term, Safety and Efficacy Study of Intranasal Esketamine in Treatment-resistant Depression”. Principal Investigator on “The Effects of Psilocybin on Cognitive Function in Healthy Participants”. Principal Investigator on “The Safety and Efficacy of Psilocybin in Participants with Treatment-Resistant Depression (P-TRD)”. UK Chief Investigator for Novartis MDD study MIJ821A12201. AY Grant funding (past and present): NIMH (USA); CIHR (Canada); NARSAD (USA); Stanley Medical Research Institute (USA); MRC (UK); Wellcome Trust (UK); Royal College of Physicians (Edin); BMA (UK); UBC-VGH Foundation (Canada); WEDC (Canada); CCS Depression Research Fund (Canada); MSFHR (Canada); NIHR (UK). Janssen (UK). No shareholdings in pharmaceutical companies.

## Funding statement

This work is funded by the European Brain Council related to the Value of Treatment research project.

## Notes

### Author Declarations

This is a scoping review protocol, which will involve human studies in the literature, for which data can be obtained by contacting the original authors where appropriate.

## References

[1] OECD and European Union. (2018). Health at a Glance: Europe 2018: State of Health in the EU Cycle. Paris/European Union, Brussels: OECD Publishing.

[2] Strawbridge, R., Zahn, R., Eberhard, J., Wasserman, D., Hegerl, U., & Brambilla, P. et al. (2021). P.0392 Care pathways for people with major depressive disorder. European Neuropsychopharmacology, 53, S281–S282. doi: 10.1016/j.euroneuro.2021.10.365

[3] Strawbridge, R., McCrone, P., Ulrichsen, A., Zahn, R., Eberhard, J., & Wasserman, D. et al. (Manuscript under review). Care pathways for people with major depressive disorder: a European Brain Council Value of Treatment study.

[4] WHO Coronavirus (COVID-19) Dashboard. (2022). Retrieved 25 January 2022, from https://covid19.who.int/

[5] World Health Organization. (2020). The impact of COVID-19 on mental, neurological and substance use services: results of a rapid assessment [Ebook]. Geneva. Retrieved from https://www.who.int/publications/i/item/978924012455

[6] OECD. (2021, May 12). Tackling the mental health impact of the COVID-19 crisis: An integrated, whole-of-society response (Policy brief). Retrieved from https://www.oecd.org/coronavirus/policy-responses/tackling-the-mental-health-impact-of-the-covid-19-crisis-an-integrated-whole-of-society-response-0ccafa0b/

[7] WHO Regional Office for Europe. (2021). Mental health and covid-19. Retrieved february 07, 2022, from https://www.euro.who.int/en/health-topics/health-emergencies/coronavirus-covid-19/publications-and-technical-guidance/mental-health-and-covid-19

[8] Weaver, N., Coffey, M., &amp; Hewitt, J. (2017). Concepts, models and measurement of continuity of care in mental health services: A systematic appraisal of the literature. Journal of Psychiatric and Mental Health Nursing, 24(6), 431–450. doi:10.1111/jpm.12387

[9] Joyce, A. S., Wild, T. C., Adair, C. E., McDougall, G. M., Gordon, A., Costigan, N., … Barnes, F. (2004). Continuity of Care in mental health services: Toward clarifying the construct. The Canadian Journal of Psychiatry, 49(8), 539–550. doi:10.1177/070674370404900805

[10] Mishra, V., Seyedzenouzi, G., Almohtadi, A., Chowdhury, T., Khashkhusha, A., Axiaq, A., Wong, W., & Harky, A. (2021). Health Inequalities During COVID-19 and Their Effects on Morbidity and Mortality. Journal of healthcare leadership, 13, 19–26. https://doi.org/10.2147/JHL.S270175

[11] Xiong, J., Lipsitz, O., Nasri, F., Lui, L., Gill, H., Phan, L., Chen-Li, D., Iacobucci, M., Ho, R., Majeed, A., & McIntyre, R. S. (2020). Impact of COVID-19 pandemic on mental health in the general population: A systematic review. Journal of affective disorders, 277, 55–64. https://doi.org/10.1016/j.jad.2020.08.001

[12] Lal, A., Erondu, N., Heymann, D., Gitahi, G., & Yates, R. (2021). Fragmented health systems in COVID-19: rectifying the misalignment between global health security and universal health coverage. The Lancet, 397(10268), 61–67. doi: 10.1016/s0140-6736(20)32228-5

[13] Nimako, K., & Kruk, M. E. (2021). Seizing the moment to rethink health systems. The Lancet. Global health, 9(12), e1758–e1762. https://doi.org/10.1016/S2214-109X(21)00356-9

[14] Tricco, A., Lillie, E., Zarin, W., O’Brien, K., Colquhoun, H., & Levac, D. et al. (2018). PRISMA Extension for Scoping Reviews (PRISMA-ScR): Checklist and Explanation. Annals Of Internal Medicine, 169(7), 467–473. doi: 10.7326/m18-0850

[15] Patel, R., Irving, J., Brinn, A., Broadbent, M., Shetty, H., & Pritchard, M. et al. (2021). Impact of the COVID-19 pandemic on remote mental healthcare and prescribing in psychiatry: an electronic health record study. BMJ Open, 11(3), e046365. doi: 10.1136/bmjopen-2020-046365

